# ACE2 Expression is Increased in the Lungs of Patients with Comorbidities Associated with Severe COVID-19

**DOI:** 10.1101/2020.03.21.20040261

**Authors:** Bruna G.G. Pinto, Antonio E.R. Oliveira, Youvika Singh, Leandro Jimenez, Andre N A. Gonçalves, Rodrigo L.T. Ogava, Rachel Creighton, Jean Pierre Schatzmann Peron, Helder I. Nakaya

**Author notes:** **Corresponding author Corresponding author information:** Helder I Nakaya, PhD, Av. Prof. Lúcio Martins Rodrigues, 370, block C, 4th floor, São Paulo – SP - Brazil, CEP: 05508-020.

## Abstract

The pandemic caused by the Severe Acute Respiratory Syndrome Coronavirus 2 (SARS-CoV-2) has resulted in several thousand deaths worldwide in just a few months. Patients who died from Coronavirus disease 2019 (COVID-19) often had comorbidities, such as hypertension, diabetes, and chronic obstructive lung disease. The angiotensin-converting enzyme 2 (ACE2) was identified as a crucial factor that facilitates SARS-CoV2 to bind and enter host cells. To date, no study has assessed the ACE2 expression in the lungs of patients with these diseases. Here, we analyzed over 700 lung transcriptome samples of patients with comorbidities associated with severe COVID-19 and found that ACE2 was highly expressed in these patients, compared to control individuals. This finding suggests that patients with such comorbidities may have higher chances of developing severe COVID-19. We also found other genes, such as *RAB1A*, that can be important for SARS-CoV-2 infection in the lung. Correlation and network analyses revealed many potential regulators of ACE2 in the human lung, including genes related to histone modifications, such as HAT1, HDAC2, and KDM5B. In fact, epigenetic marks found in ACE2 locus were compatible to with those promoted by KDM5B. Our systems biology approach offers a possible explanation for increase of COVID-19 severity in patients with certain comorbidities.

## Introduction

Recent studies of the epidemiological characteristics of COVID-19 have revealed that severe infection is more likely in people with an existing chronic medical condition. Two independent studies of infected populations in Wuhan, China found that approximately half the subjects infected with COVID-19 had an existing comorbidity [1, 2]. In a study of 1099 patients across mainland China, 38.7% of patients with comorbidities progressed to severe infection [3]. 2020), and in a study of 52 inpatients in Wuhan, 67% of patients with comorbidities died [2]. The most common comorbidities reported in these studies were hypertension, diabetes, cerebrovascular disease, chronic obstructive lung disease, and coronary heart disease [1-3]. Other comorbidities such as carcinoma, chronic kidney disease, chronic liver disease, digestive system disease, and nervous system disease have also been reported in patients with COVID-19 [1, 2, 4]. A better understanding of the link between these conditions and COVID-19 infection is required to inform better treatment and prevention interventions.

The molecular mechanism responsible for the increased disease severity in patients with these comorbidities is not fully understood, but previous studies suggest a role for angiotensin-converting enzyme 2 (ACE2) [5]. ACE2 is a membrane protein required for SARS-CoV2 to bind and enter cells [6-8]. After binding, viral entry is facilitated by the activation of the viral spike glycoprotein and cleavage of the C-terminal segment of ACE2 by proteases like TMPRSS2 and FURIN that are readily expressed in lung tissue [9-11]. ACE2 is only moderately expressed in healthy lung tissue compared to the heart, kidneys, and testes [12], but staining of lung tissue sections from adults with pulmonary hypertension has revealed increased ACE2 protein in the endothelium of pulmonary arteries, compared to healthy controls [13]. ACE2 upregulation has also been observed in animal models of liver fibrosis [14]. However, the reason for this upregulation remains unclear, and a link to other COVID-19 comorbidities has not been determined.

Here, we showed that ACE2 expression in lung tissue is upregulated by diseases representing comorbidities along with COVID-19. We also used systems biology approaches including co-expression analysis, meta-analysis, and network analysis to determine a potential cause of the ACE2 upregulation. From this analysis, we found that ACE2 expression could be regulated by enzymes that modify histones, including KDM5B. This identification of a common molecular mechanism of increased COVID-19 severity in patients with diverse comorbidities could direct the development of interventions to reduce the infection risk and disease severity in this population.

## Methods

### Literature curation

Relevant scientific literature related to key COVID-19 morbidities was retrieved from PubMed. For terms returning more than 100,000 papers (“hypertension,” “smoking,” and “asthma”), only the most recent 100,000 papers were analyzed. Full papers and abstracts were annotated to identify all genes, diseases, and species appearing in the title or abstract using the PubTator Central API [15]. This open source tool uses TaggerOne for disease annotations, GNormPlus for gene annotations, and SR4GN for species annotations [15]. The data were filtered to retain only papers containing a human species annotation. Next, every possible combination of gene and disease annotation within the title and abstract of each paper was generated. Only gene-disease terms with at least four documents supporting it, and those with a proximity less than or equal to the median sentence length of the paper section were retained.

Gene IDs were converted to gene symbols using the biomaRt R package [16, 17], and disease IDs were converted to disease MeSH terms using the Entrez Programming Utilities to query the Entrez database provided by the National Center for Biotechnology Information. The data was then further filtered to retain disease MeSH terms relevant to reported clinical COVID-19 comorbidities [3]. Redundant terms were collapsed using fuzzy string matching. The final gene-disease data set was used to generate a network utilizing Gephi software where the nodes were genes and diseases, and the edge weight was determined by the number of analyzed papers containing the gene-disease combination [18].

### Meta-Analysis

We manually curated Gene Expression Omnibus (GEO) repository (https://www.ncbi.nlm.nih.gov/geo/) to find lung and blood transcriptome datasets related to “Pulmonary Arterial Hypertension” (PAH), “Chronic Obstructive Pulmonary Disease” (COPD), “Severe Acute Respiratory Syndrome” (SARS), “H1N1 Influenza”, “Tuberculosis” and “Smoking”. Author-normalized expression values and metadata from these datasets were downloaded using the GEOquery package [19]. We performed differential expression analyses between patients with a disease and the control individuals (see Table S1) using the limma package [20]. The gene symbol for each probe was obtained from the annotation file [21]. Probes that matched the same gene symbol were collapsed by taking the one with the lowest p-value. Meta-analysis was performed with the MetaVolcanoR package [22] by combining the p-values using Fisher’s method. We calculated the False Discovery Rate (FDR) to identify the differentially expressed genes (FDR < 0.05). For enrichment analyses, we utilized the EnrichR tool [23] with the “GO Biological Process 2018” and “BioPlanet 2019” databases. We then selected pathways with an Adjusted P-value < 0.05. The network was created in Cytoscape [24].

## Results

To identify the genes highly associated with key comorbidities of severe COVID-19 [1, 3], we mined all relevant scientific literature of these human diseases. Specifically, over 8,000 abstracts containing the terms “pulmonary hypertension,” “chronic obstructive pulmonary disease,” “hypertension,” “smoking,” “pulmonary fibrosis,” or “asthma” in the title or abstract were selected (Figure 1a). Our text-mining analysis revealed 804 genes highly associated with one or more COVID-19 morbidities (Figure 1b). Among those genes, 26 were associated with four or more diseases (Figure 1c). Although ACE2 was known to be related to “Cardiovascular Diseases”, “Familial Primary Pulmonary Hypertension”, “Hypertension, Pulmonary”, and “Hypertension”, none of the articles containing this gene-disease association studied how ACE2 expression was altered in the lungs of patients with these diseases.

**Figure 1.**
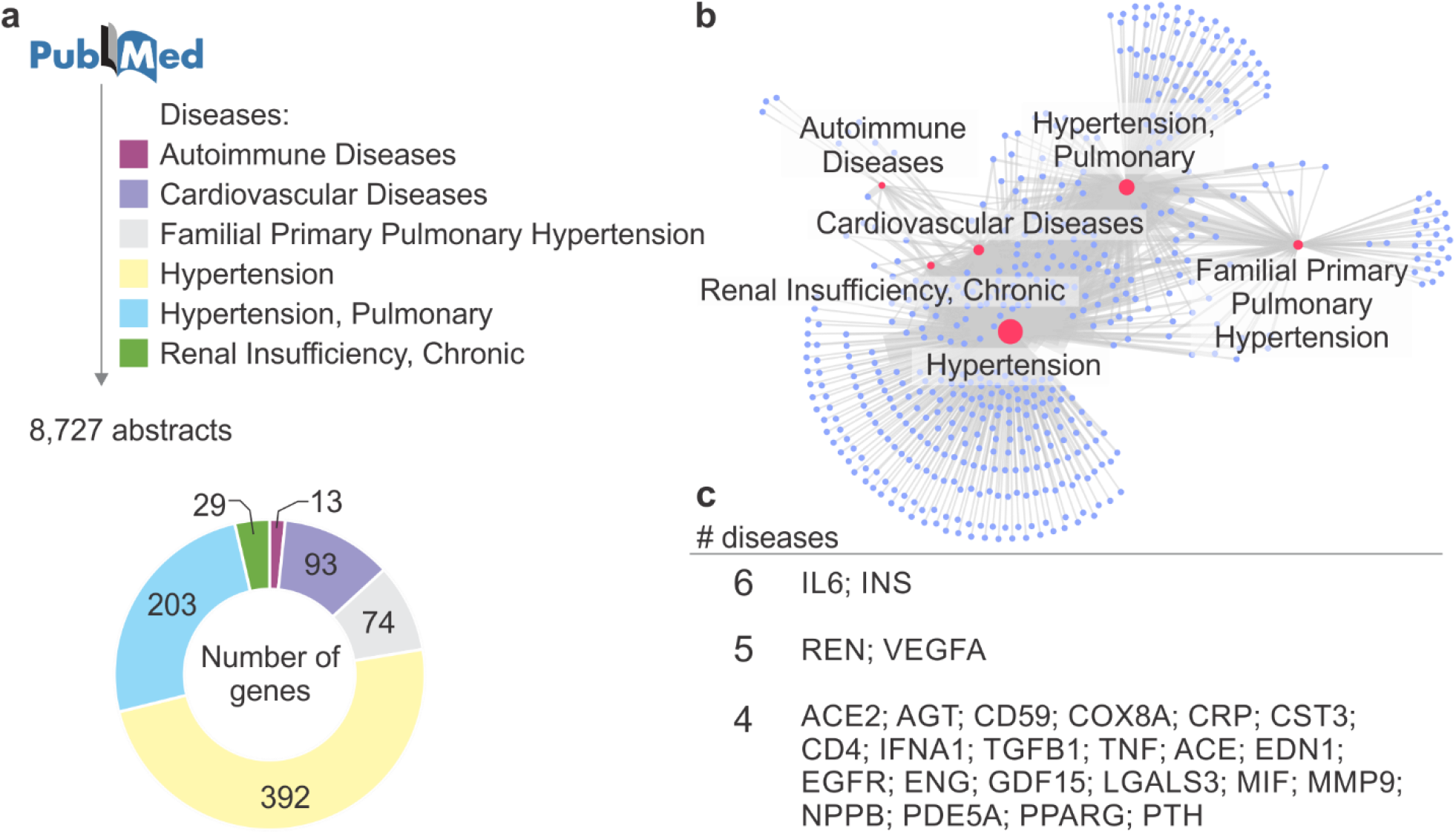
Literature curation of genes associated with key COVID-19 morbidities. **a**. Text-mining approach to retrieve genes in abstracts associated with six human diseases. The number of genes present in at least four abstracts of a disease is shown in the pie chart. **b**. The knowledge-based network of COVID-19 morbidities. The network shows the diseases (red nodes) and genes (purple nodes) from panel a. The edges represent an association between a disease and a gene. The size of the nodes is proportional to its degree. **c**. Genes associated with four or more COVID-19 morbidities.

Seven lung transcriptome studies of patients with either Chronic Obstructive Pulmonary Disease (COPD) or Pulmonary Arterial Hypertension (PAH), as well as smoking volunteers, compared to individuals who were non-smoking volunteers, were downloaded and used in our meta-analysis (Table S1). For each study, we performed differential expression analysis between patients and control individuals (Table S1). By combining the p-values obtained in all the seven comparisons, we were able to identify 1,740 and 938 genes that were, respectively, up- and down-regulated in the disease (Figure 2a). Enrichment analysis using these differentially expressed genes revealed several pathways associated with inflammatory processes, metabolism, and ER stress. Among the pathways enriched with down-regulated genes, there were “vasculogenesis” and “regulation of Notch signaling pathway” (figure 2b). The “viral life cycle” pathway, which describes the processes utilized by viruses to ensure survival and to attach and enter the host cells was enriched with up-regulated genes (Figure 2b). ACE2 was included in this pathway, as well as 25 other genes (Figure 2c). One of these genes is *RAB1A*. Rab GTPases are involved in the replication of many viruses infecting humans [25], but have not been associated with SARS-CoV-2 life cycle yet. Both *TMPRSS2* gene, which is required for SARS-CoV-2 cell entry [5], and *FURIN* gene, which cleaves SARS-CoV-2 spike glycoprotein [26] were not differentially expressed in most of the lung transcriptome. However, both genes were highly expressed in lung (data not shown), suggesting that the levels of ACE2 may be the limiting factor for viral infection.

**Figure 2.**
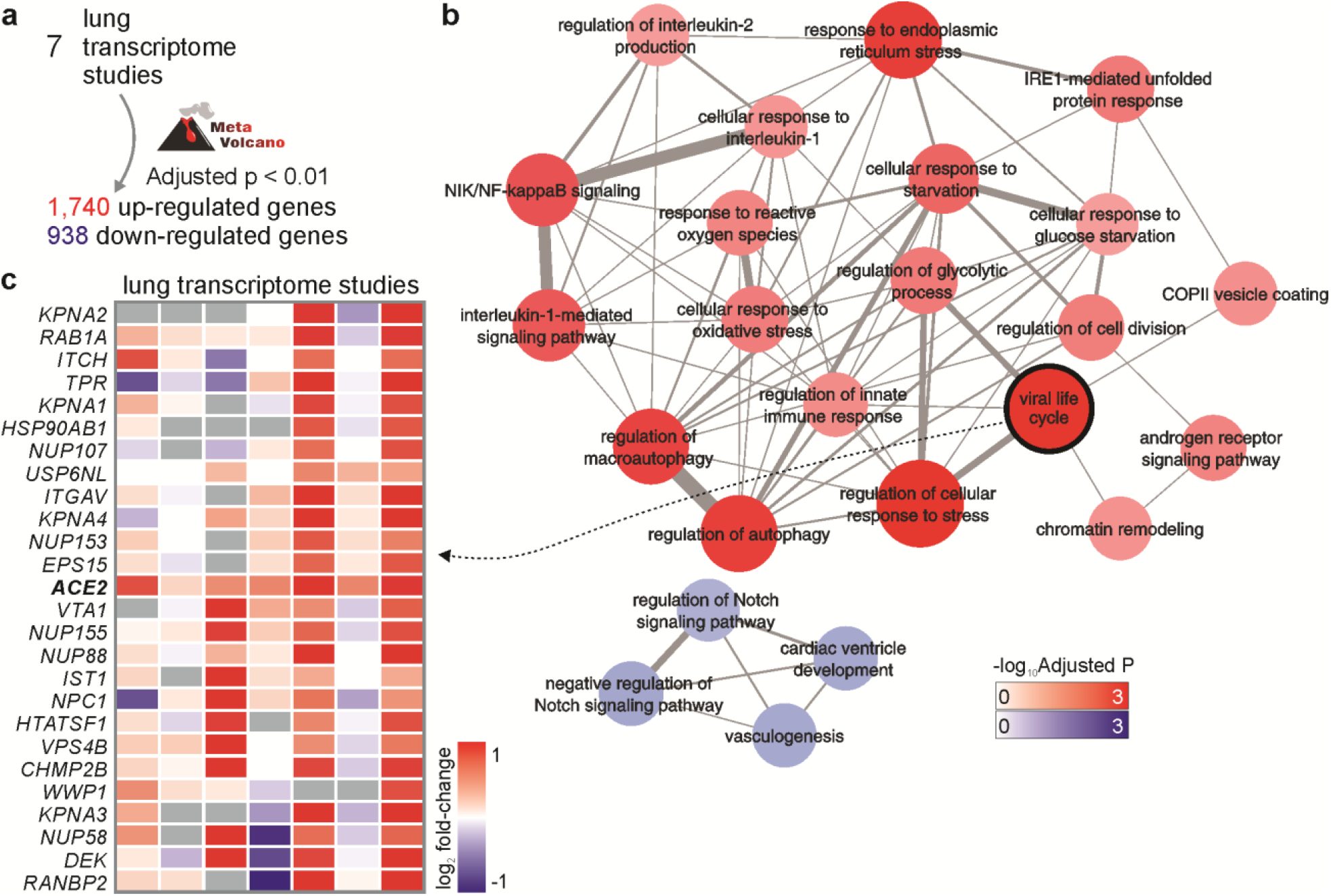
Meta-analysis of lung transcriptomes of patients with COVID-19 morbidities. **a**. Meta-analysis of seven differential expression analyses. Meta-volcano tool was used to combine the p-values of seven studies (Table S1) and to identify the differentially expressed genes (FDR < 0.01). **b**. Pathway enrichment analysis. Pathways from the “GO Biological Process 2018” database with Adjusted P-value < 0.05 were selected to create the network. The width of edges is proportional to the number of genes shared by two pathways (nodes). The size and color of nodes are proportional to the - log10 Adjusted P-value. **c**. Genes from the “viral life cycle” pathway that were up-regulated in human diseases. The colors in the heat map represent the log2 fold-change between patients and control individuals.

We then decided to investigate whether ACE2 was specifically up-regulated in the lungs of patients having one of these morbidities (Figure 3a). Since cancer is also listed as a relevant co-morbidity [3], we analyzed an RNA-seq data of patients with lung adenocarcinoma [27]. ACE2 was expressed in 59% of cancer samples and only in 25% of adjacent lung normal samples (Figure 3b). Among the samples expressing ACE2, the level of the gene was higher in cancer, compared to the adjacent normal lung tissue (Figure 3b). In another lung RNA-seq dataset, we compared ACE2 expression between patients with COPD and subjects with normal spirometry [28]. Again, the expression of ACE2 was significantly up-regulated in the disease compared to controls (Figure 3c). In fact, ACE2 was significantly up-regulated in 6 out of 7 lung transcriptome studies (Figure 3d-e), suggesting that patients who have COPD or PAH, and even people who smoke, may have higher chances of developing severe COVID-19.

**Figure 3.**
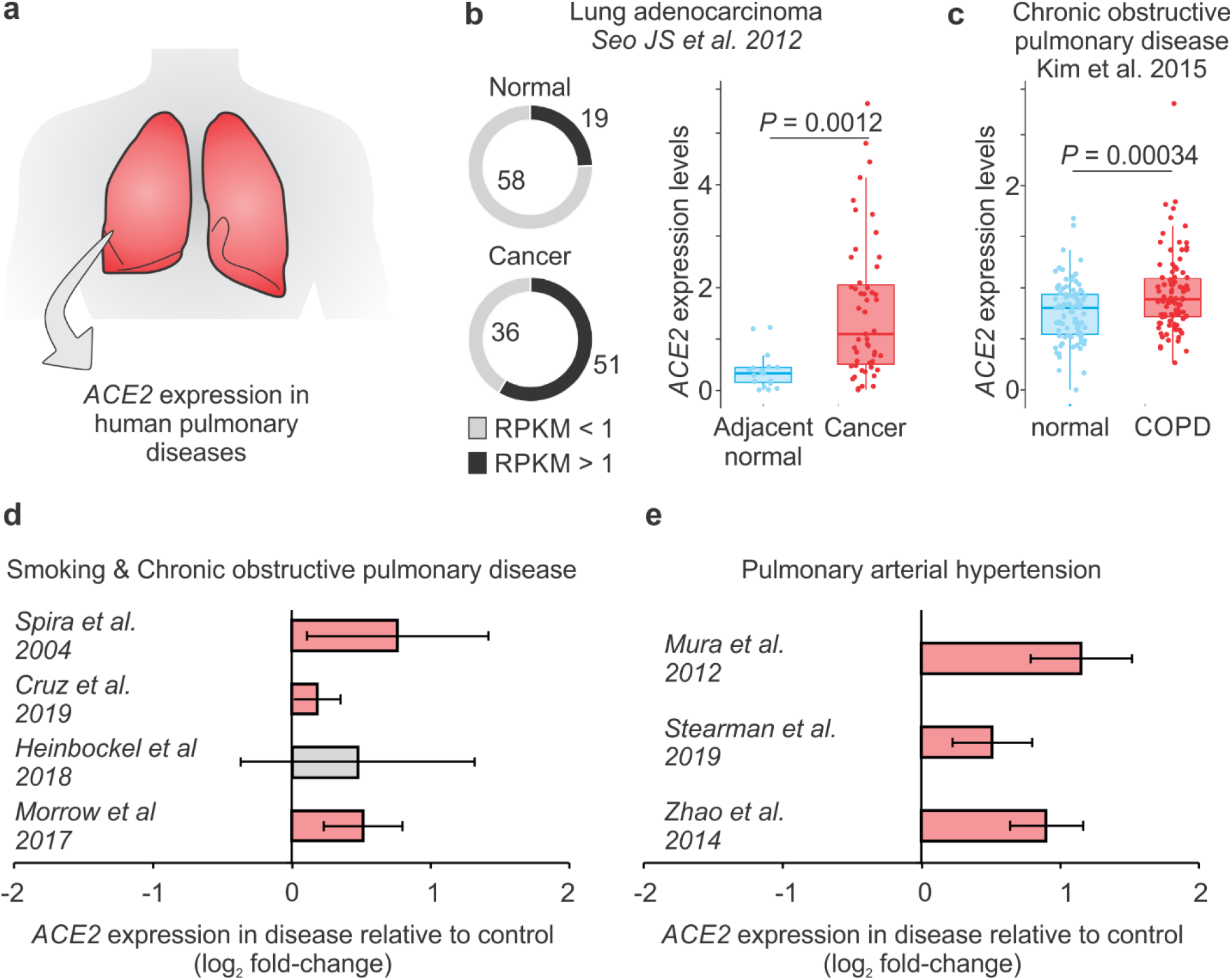
ACE2 is up-regulated in patients with lung diseases. **a**. Analysis of ACE2 expression in lung transcriptome datasets of patients with human pulmonary diseases. **b**. ACE2 expression in patients with lung adenocarcinoma. The pie chart in the left shows the number of samples with (black) or without (grey) ACE2 expression. RPKM: Reads Per Kilobase of transcript, per Million mapped reads. The boxplot in the right shows the difference between cancer cells (red dots) and adjacent normal cells (blue dots). Student t-test P-value is indicated. **c**. ACE2 expression in patients with COPD. The boxplot on the right shows the difference between COPD patients (red dots) and control individuals (blue dots). Student t-test P-value is indicated. **d**. ACE2 is up-regulated in patients with COVID-19 morbidities. Each bar represents the log2 expression fold-change between patients and control individuals. The error bars indicate the 95% confidence interval. Bars in red represent a p-value < 0.05 and in grey a non-significant p-value. The original studies are indicated and can be found in Table S1.

Co-expression analyses can provide useful insights about the functional role of genes and their regulatory mechanisms [29]. We performed Pearson correlation between the expression of ACE2 and all other genes in each of the seven lung transcriptome studies (Table S1), combined the p-values using Fisher’s method, and applied an FDR correction (Figure 4a). This approach identified 544 and 173 genes with positive and negative correlation with ACE2, respectively (Figure 4a). Several of these genes were related to histone modifications, such as HAT1, HDAC2, KDM5B, among others (Figure 4a). Among the positively correlated genes, we found ADAM10 that regulates ACE2 cleavage in human airway epithelia [30] and TLR3 that plays a key role in the innate response to SARS-CoV or MERS-CoV infection [31].

**Figure 4.**
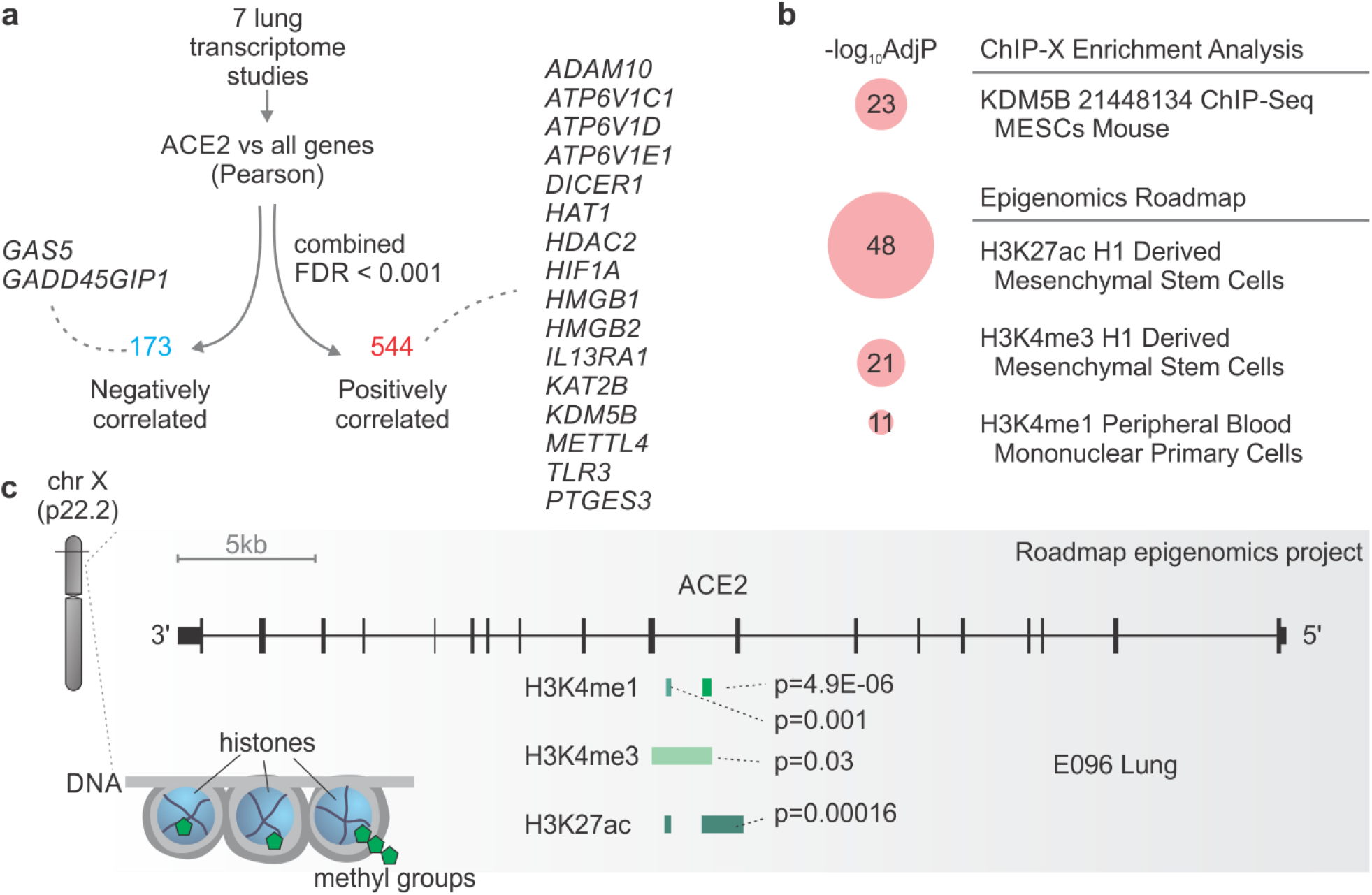
Insights of ACE2 regulation in the lung. **a**. Genes whose expression is correlated with ACE2 in the lung. Selected genes that were negatively (blue) or positively (red) correlated with ACE2 are highlighted. **b**. Pathway enrichment analysis using the ACE2-positively correlated genes. Pathways from the “ChIP-X Enrichment Analysis” and “Epigenomics Roadmap” databases with Adjusted P-value < 10^−10^ were selected. The size of the red circles is proportional to the - log_10_ Adjusted P-value of the enrichment. **c**. ACE2 locus contains marks of histone acetylation and methylation. The plot was modified from the WashU EpiGenome Browser using E096 lung. The peaks corresponding to each histone modification and the p-values of the marks are indicated.

Pathway enrichment analysis revealed that several of the genes positively associated with ACE2 were regulated by KDM5B, and by specific histone acetylation (H3K27ac) and histone methylation (H3K4me1 and H3K4me3) (Figure 4b). In fact, KMD5B demethylates lysine 4 of histone H3 (i.e. H3K4) and is involved in transcriptional regulation and DNA repair [32]. We then checked in the Roadmap Epigenomics Project database [33] to see whether ACE2 locus contained ChIP-seq information for these histone markers. In the human lung, peaks for H3K4me1 and H3K4me3, as well as H3K27ac, were identified in ACE2 locus (Figure 4c), suggesting that ACE2 may be epigenetically regulated in the lung.

## Discussion

The current diabetes pandemic [34] could be worsening the SARS-CoV-2 pandemic by increasing the comorbidities associated with severe COVID-19. As we did not find lung transcriptome samples from patients with type 2 diabetes, we could not directly test whether ACE2 expression was increased in patients with diabetes, compared to healthy controls. However, our text-mining approach revealed that IL-6 and INS genes were associated with all the diseases we searched. The *INS* gene encodes the insulin hormone, and insulin is associated with the NAD-dependent histone deacetylase Sirtuin 1 (SIRT1) [35]. We found that SIRT1 was up-regulated in the lung of patients with severe COVID-19 comorbidities in 4 out 7 studies (data not shown). Clarke et al [36] have demonstrated that, under conditions of cell energy stress, SIRT1 can epigenetically regulate ACE2. Others too have shown that non-steroidal anti-inflammatory drugs may inhibit the SIRT1 deacetylase activity [37], which in turn could impact ACE2 expression.

The “viral life cycle” pathway that was enriched with up-regulated genes in patients with severe COVID-19 comorbidities contains several genes other than ACE2 that can be potentially important for SARS-CoV-2 cell cycle and invasion/attachment. These include *RAB1A* gene, whose product promotes the replication of Vaccinia virus [38]. Also, RAB1A is important for Herpes Simplex Virus 1 Secondary Envelopment [39], and is required for assembly of classical swine fever virus particle [40]. It is possible that SARS-CoV-2 utilizes RAB1A as well.

The fact that ACE2 gene is located in the X chromosome, and the initial findings showing that older males with comorbidities are more likely to be have severe COVID-19 compared to females [1], indicate that ACE2 expression in the lung may be sex-biased. Although no significant sexual differences was found in the activity of ACE2 in mouse lung [41], in rats, the levels of ACE2 were dramatically reduced with aging in both genders, but with significantly higher ACE2 expression in old female rats than male [42].

Although the mechanisms by which *ACE2* is up-regulated in patients with severe COVID-19 comorbidities were not addressed, our analysis may shed some light on the subject. Among the genes whose expression was positively correlated with ACE2, we detected genes associated with epigenetic regulation of gene transcription. For instance, HAT and HDAC modulate chromatin and DNA condensation by changing histone acetylation status, thus permitting gene transcription. This could be happening in lung tissue, facilitating ACE2 expression, as observed during lung cancer and COPD.

KDM5B is associated with infection of hepatitis B virus [43]. In breast cancer cells, blockage of KDM5 triggers a robust interferon response that results in resistance to infection by DNA and RNA viruses [44]. This finding suggests that KDM5 demethylases are potential targets for preventing SARS-CoV-2 infection.

COVID-19 may kill between 5.6% and 15.2% of people infected with SARS-CoV-2 [45]. Drug treatments that lower this mortality rate may save many thousands of lives. Our systems biology approach offers putative gene targets for treating and preventing severe COVID-19 cases.

## Data Availability

All data can be found in Table S1.

## Acknowledgments

We would like to thank Tiago Lubiana for his valuable inputs.

## Funding

HIN is supported by CNPq (313662/2017-7) and the São Paulo Research Foundation (FAPESP; grants 2017/50137-3, 2012/19278-6, 2018/14933-2, 2018/21934-5 and 2013/08216-2). RC was supported by an NSF Graduate Research Fellowship (DGE-1256082), NSF Graduate Research Opportunities Worldwide Fellowship, and by the STD & AIDS Research Training Fellowship Program (NIH 2T32AI007140-41).

## Author contributions

BGGP, AERO, YS, LJ, ANAG, RLTO, RC, HIN performed the analyses. JPSP and HIN interpreted the results. All authors helped with the writing of the manuscript.

## Declaration of Interest

The author reports no conflicts of interest in this work.

## Ethical Approval

Not applicable as we utilized publicly available data.

## Notes

### Competing Interest Statement

The authors have declared no competing interest.

